# Associations between exposure to single cadmium, lead, mercury and mixtures and women’s infertility and long-term amenorrhea

**DOI:** 10.1101/2022.10.31.22281773

**Authors:** Maria McClam, Jihong Liu, Yihan Fan, Tingjie Zhan, Qiang Zhang, Dwayne E. Porter, Geoffrey I. Scott, Shuo Xiao

## Abstract

**Purpose:** Cadmium (Cd), lead (Pb), and mercury (Hg) have been shown to exhibit endocrine disrupting properties. their effects on women’s reproductive health, however, remain elusive. Here, we investigated associations between blood concentrations of single of Pb, Cd, Hg, and their mixture and infertility and long-term amenorrhea in women of reproductive age using the US National Health and Nutrition Examination Survey (NHANES) 2013-2018 cross-sectional survey.

**Methods:** A total of 1,990 women were included for the analysis of infertility and 1,919 women for long-term amenorrhea. The methods of log-transformation and quarterization were used to analyze blood heavy metal concentrations. Statistical differences in the covariates between the outcome groups were evaluated using a chi-squared test for categorical variables and a t-test for continuous variables. Multiple logistic regression models were used to examine the associations.

**Results:** The blood concentrations of Pb and heavy metal mixtures were significantly higher in ever-infertile women than pregnant women, but the concentrations of Cd and Hg were comparable. Multiple logistic regression analysis revealed that after the full adjustment, there was a significant and dose-dependent positive association between blood Pb concentrations and women’s historical infertility, a negative association between Cd and women’s long-term amenorrhea, and no associations between Hg and heavy metal mixture and women’s infertility or long-term amenorrhea.

**Conclusions:** Our study demonstrates that exposure to heavy metals exhibit differential associations with women’s infertility and long-term amenorrhea.

**Disclosure summary:** The authors declare no conflict of interest.

## Introduction

The female reproductive system provides hormonal control and anatomical structure to sustain a woman’s menstrual cycle and fertility. Infertility is the failure of achieving clinical pregnancy after one year of unprotected intercourse, affecting up to 15% of couples worldwide.^1,2^ In the US, the number of women with impaired fertility has been estimated to increase from 4.5 million in the early 1980s to about 7.7 million by 2025.^3^ Although women’s infertility can be caused by male factors and unexplained reasons,^4^ the majority of them have recognized reproductive or neuroendocrine disorders, such as premature ovarian insufficiency (POI),^5^ oligomenorrhea or amenorrhea,^1^ anovulation, ^6^ poor gamete quality,^7^ and other reproductive diseases such as polycystic ovarian syndrome (PCOS),^8^ endometriosis,^9^ and hypothalamic dysfunction.^10^ So far, the mechanism of women’s infertility remains incompletely understood but has been attributed to both genetic factors and exposure to reproductive toxicants.^11^

Industrial development, agricultural practices, and the production and use of consumer products have introduced various toxic substances into the environment, including heavy metals that are naturally occurring metallic elements with high molecular weight and density.^12^ Cadmium (Cd), lead (Pb), and mercury (Hg) are three primary heavy metals listed by the World Health Organization (WHO) under the top 10 toxicants of major public health concern.^13^ The environmental contamination of heavy metals primarily stems from industrial mining, agricultural practice, and fossil fuel and waste combustion, etc.^14-17^ Heavy metals persist and bioaccumulate along the food chain and in drinking water, soils, and air, making them a major source of environmental toxicants to humans.^18^

Women’s reproductive health is vulnerable to environmental toxins, particularly endocrine disrupting chemicals (EDCs) that interfere with the body’s normal hormone synthesis, secretion, and signaling.^19,20^ Growing epidemiological and experimental research have revealed that heavy metals exert endocrine disrupting properties,^21-25^ implicating the possible causative relationship between exposure to heavy metals and women’s infertility and other reproductive disorders. In a cross-sectional study that compared 310 women with clinically diagnosed infertility and 57 pregnant women in Taiwan, the blood concentrations of Pb but not Cd in infertile women were significantly higher than pregnant women.^26^ Another study compared 82 infertile and 42 pregnant women in the US and found that there were positive associations between blood concentrations of Pb and Cd and women’s infertility.^27^ Heavy metals have also been shown to affect reproductive hormone secretion. In premenopausal women, the blood concentrations of Cd, Pb, and Hg were associated with altered means and amplitudes of follicle stimulating hormone (FSH) and luteinizing hormone (LH), two gonadotropins that regulate ovarian follicle maturation, hormone secretion, and ovulation.^28^ It was also found in the same study that Pb may increase progesterone levels in the follicular phase, and both Pb and Hg cause a delay of the progesterone rise in the mid-luteal phase.^28^

Experimental research has documented that exposure to heavy metals may impact the female reproductive cycle and fertility. For example, Cd exposure in mice compromised oocyte meiotic and developmental competence by inducing oocyte oxidative stress, early apoptosis, and epigenetic modifications, which eventually resulted in decreases in litter size.^29^ Pb has been found to delay vaginal opening, decrease estradiol secretion, and interfere with ovarian cyclicity in rats, suggesting the harmful effects of Pb on the ovaries or the entire hypothalamic-pituitary-gonadal (HPG) axis.^30^ Heavy metals may also act as agonists or antagonists to disrupt hormone receptor-mediated signaling. All Cd, Pb, and Hg have been reported to exert estrogenic effects by binding to the estrogen receptor α and/or β, which may disrupt the expression of estrogen target genes and the proliferation and/or differentiation of estrogen-responsive tissues such as the endometrium.^23,24^ Altogether, existing epidemiological and experimental evidence suggests that exposure to heavy metals may perturb women’s menstrual cycle and fertility by interfering with the homeostasis of the HPG axis, ovarian steroidogenesis, hormonal signaling, and other reproductive events. However, the majority of the epidemiological studies have small sample sizes and do not consider the complexities of the female reproductive cycle and fertility;^26-28,31^ moreover, previous studies primarily focused on a single metal at a time, but women are periodically or even constantly exposed to mixtures of multiple heavy metals, which may cause cumulative effects.^26-28,31-33^

The objective of this study is to investigate associations between blood concentrations of single Pb, Cd, Hg and their mixtures and reproductive aged women’s infertility in the National Health and Nutrition Examination Survey (NHANES) 2013-2018; moreover, the associations between heavy metals and women’s long-term amenorrhea, a crucial contributing factor to women’s infertility, was assessed. We hypothesize that women with higher blood heavy metal concentrations are more likely to experience infertility and long-term amenorrhea. We combined our robust understanding of female reproductive biology and epidemiology to create a comprehensive evaluation of the impacts of exposure to single heavy metals and their mixtures on women’s reproductive health.

## Materials and Methods

### Study population

All data were obtained from NHANES, a nationally representative cross-sectional survey of the non-institutionalized U.S. population. NHANES is conducted by the US Centers for Disease Control and Prevention (CDC) and uses a complex multistage, probability sampling design. Since 1999, the sample design has consisted of multi-year, stratified, clustered four-stage samples, with data released in 2-year cycles. NHANES samples are drawn in four stages: (1) Primary sampling units (PSUs) (counties, clusters of tracts within counties, or combinations of neighboring counties), (2) segments within PSUs (census blocks or groupings of blocks), (3) dwelling units (DUs) (households) within segments, and (4) individuals within households. Screening is conducted at the DU level to identify individuals, based on oversampling criteria. NHANES oversamples some subgroups to increase the reliability and precision of health status indicator estimates for these particular subgroups; the population subgroups chosen for oversampling directly determine the sampling domains used to select the sample at all stages.^34^ In this study, we used data from three continuous NHANES cycles, including 2013-2014, 2015-2016, and 2017-2018, where the reproductive health questionnaire addressed women’s infertility and menstrual cycle. All data including sociodemographic questionnaires, physical examinations, and reproductive health questionnaires, were downloaded directly from the CDC’s website.^35^

### Study sample, variable descriptions, and inclusion

Among all three NHANES cycles, one-half of participants age 12+ have blood heavy metal data for the cycles of 2013-2014 and 2015-2016. All participants aged 1+ have blood heavy metal data available for the cycle of 2017-2018. The total number of participants in these three NHANES cycles was 20,113. After excluding males (n=9,934), females younger than 20 years (n=4,589), and females older than 49 years (n=2,843), there were 2,747 reproductive aged women (20-49 years) who had blood heavy metal data available. Although post-pubertal women under 20 years are also considered within reproductive age, they were not included because NHANES survey was designed to only collect reproductive data from participants 20 years of age and older. Moreover, women who had a hysterectomy (n=125) and women with missing data for the heavy metal exposures (n=136) were also excluded. Figures 1 and 2 describe the sample attrition process and amount of missingness. Participants with missing data for the questions of infertility (n=272), demographic variables (n=197), BMI (n=14), and information on the use of birth control pill and female hormones (n=4) were also excluded. Overall, a total of 1,999 women were included for comparing ever-infertile and fertile women (main group), and a total of 297 participants were included for comparing ever-infertile and pregnant women (sub-group) (Figure 1). For assessing long-term amenorrhea, participants with missing data for the questions of long-term amenorrhea (n=361), demographic variables (n=190), BMI (n=12), and information on the use of birth control pill and female hormone use (n=4) were also excluded. Overall, a total of 1,919 women were included for assessing long-term amenorrhea (Figure 2).

**Figure 1:**
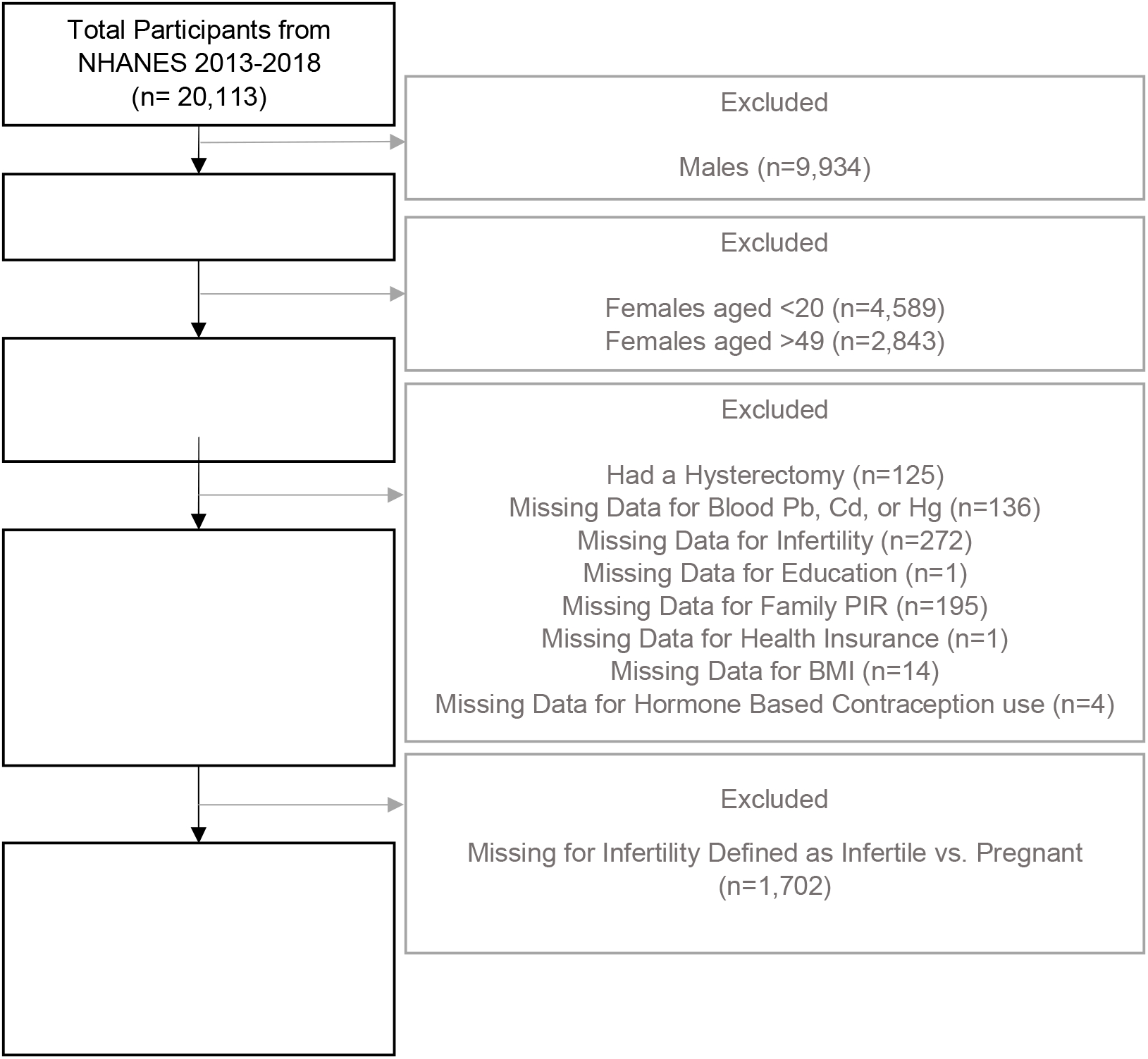
Schematic diagram depicting the process of inclusion of women from NHANES 2013–2018 for investigating associations between blood heavy metal concentrations and women’s fertility.

**Figure 2:**
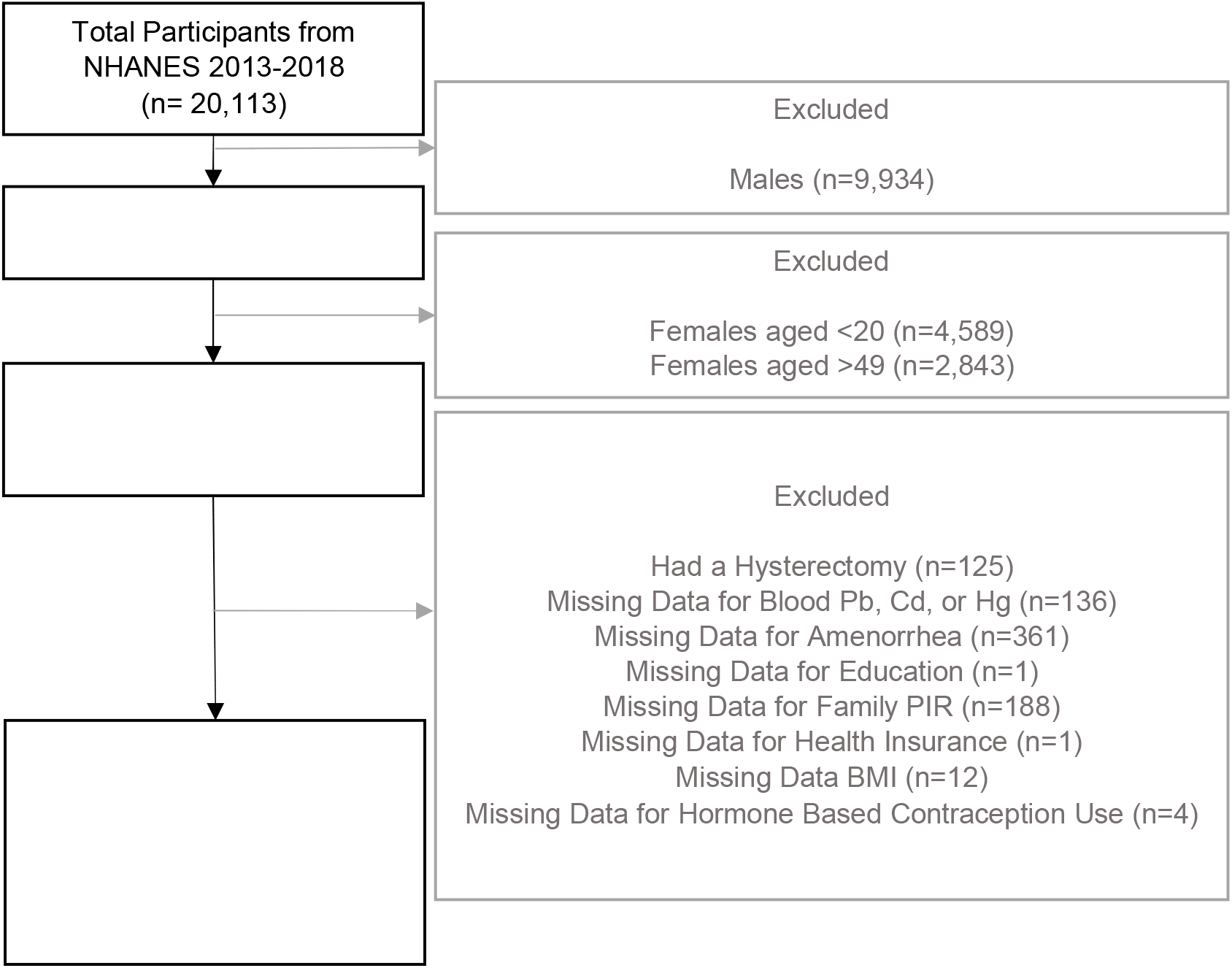
Schematic diagram depicting the process of inclusion of women from NHANES 2013–2018 for investigating associations between blood heavy metal concentrations and women’s long-term amenorrhea.

### Measurements of blood Pb, Cd, Hg concentrations

The blood concentrations of Pb, Cd, and Hg were measured in the whole blood using the mass spectrometry after a simple dilution sample preparation step. The full NHANES laboratory procedures can be found online.^36-38^ The lower limit of detection (LLOD) of the three measured metals were: 0.07 μg/dL for Pb, 0.1 μg/dL for Cd, and 0.28 μg/dL for Hg. For analytes with analytic results below the LLOD, an imputed fill value was placed in the analyte results field. This value is LLOD divided by the square root of 2 (LLOD/sqrt [2]).

### Creating a metal mixture value

Previous studies have used simple additive methods by summing all metal scores with equal weight to create a score of the metal mixture.^39,40^ Here, we aimed to further fine tune this mixed metal score by using a novel method, toxic equivalency (TEQ) values that are a weighted quantity measure based on the relative toxicity potency of each chemical. TEQ values are used for reporting dioxin and dioxin-like compounds.^41^ We used a similar methodology to create TEQ values for the mixture of the three heavy metals. Pb, Cd, and Hg have been shown to exhibit similar toxic mechanisms by inducing oxidative stress and endoplasmic reticulum (ER) stress,^42,43^ which compromises the reduction-oxidation hemostasis and eventually results in adverse health outcomes.^44-46^ ER stress has also been revealed as a key molecular mechanism in various female reproductive functions and disorders, such as ovarian injury via ER stress-mediated apoptosis/autophagy, regulation of gestational length by the uterine ER stress, oocyte maturation, and embryo implantation.^47-50^

In the federal Tox21 program, the ER Stress Response Element β-lactamase reporter gene assay (ESRE-bla) is used to screen potential toxicants, including heavy metals.^51,52^ Pb, Cd, and Hg in certain form have been shown to be ‘active’ in TOX21_ESRE_BLA assay, while other high-throughput assays related to oxidative stress lack the screening results for all three heavy metals in this study. Data from the assay component TOX21_ESRE_BLA_ratio were extracted from the CompTox Chemistry Dashboard for Lead(II) acetate trihydrate, Cadmium acetate dihydrate, and Mercury(II) acetate.^53^ The concentration of the half-maximal activity (AC50), a common potency measure applied in pharmacological research and toxicity testing^54^ was identified for each heavy metal: Lead(II) acetate trihydrate AC50 = 0.0586 μM, Cadmium acetate dihydrate AC50 = 0.0545 μM, and Mercury(II) acetate AC50 = 2.29 μM. The maximal response or efficacy of the three heavy metals are in the same order of magnitude, with that of Cd and Hg within two-fold of Pb, which is used as the reference metal to calculate the TEQ values of the other two.^55,56^ Using AC50, the adjusted metal weights were 4.831e-2 for Pb, 9.565e-3 for Cd, and 1.276e-4 for Hg. The final mixed metal score was calculated using the sum of weighted blood metal concentrations as follows: Mix Metal Score = [(1*Pb Blood Metal Concentration, μg/dL*10 / 207 g/mol) + (1.0752*Cd Blood Metal Concentration, μg/L / 112.41 g/mol) + (0.0256*Hg Blood Metal Concentration, μg/L / 200.59 g/mol)] *100. The simplified formula is [(4.831e-2*Pb Blood Metal Concentration) + (9.565e-3*Cd Blood Metal Concentration) + (1.276e-4*Hg Blood Metal Concentration)] *100. Following TEQ approach, we refer to this as our metal mixture value of exposure throughout the paper.

### Women’s infertility history

The prevalence of infertility among women aged 20-49 was assessed using the question “Have you ever attempted to become pregnant over a period of at least a year without becoming pregnant?”.^1^ Women who responded “Yes” were considered ever-infertile. Fertile women were defined in two distinct ways: (1) fertile women or the main-group were women who answered “No” to the question of “Have you ever attempted to become pregnant over a period of at least a year without becoming pregnant?”, and (2) pregnant women or the sub-group who answered “Yes” to the question “Are you pregnant now?”. Infertility defined using this method represents a women’s history of infertility and may not reflect their current fertility status; hence we also analyzed women’s recent long-term amenorrhea in this study.

### Women’s recent long-term amenorrhea

Women with long-term amenorrhea were defined by those who answered “no” to the question “Have you had at least one menstrual period in the past 12 months? (Please do not include bleedings caused by medical conditions, hormone therapy, or surgeries.)” and answered “Other” or “Don’t know” to the question “What is the reason that you have not had a period in the past 12 months?”. Menstruating women were defined by women who answered “Yes” to the same question. Participants who answered “Pregnancy”, “Breast feeding”, and “Menopause/Change of life” to the question “What is the reason that you have not had a period in the past 12 months?” were excluded from this study. The outcome variable long-term amenorrhea defined here reflects the women’s current or recent menstrual cycle status in the past 12 months. Although menopause is defined as amenorrhea for 12 consecutive months,^57^ these women did not self-report having menopause; thus, our outcome of long-term amenorrhea may reflect their most recent (last 12 months) or current fertility status.

### Other Covariates

Age was included as a covariate because age is an important factor determining a woman’s menstrual cycle, menopause, and fertility. Demographic variables including race/ethnicity, education, family poverty income ratios were all included as covariates. Because this study assessed women’s reproductive capacity, which closely ties to sexual relationships, we included marital status as a covariate. We also included health insurance coverage as a covariate because health care access can impact participants’ reproductive health and fertility management.^58^ Smoking status and BMI were included because they have been shown to impact women’s reproductive health.^59,60^ BMI was defined by the CDC as underweight (<18.5), healthy weight (18.5 to <25), overweight (25 to <30), and obesity (30 or higher).^61^ Hormonal contraception use was included because women are often prescribed hormones to regulate menstruation or prevent menstruation and unintended pregnancy. Hormonal contraception use included women who have ever taken birth control pills or used female hormones. Additionally, when assessing infertility as an outcome, two additional covariates were included: regular menstruation and if women had seen a doctor because they were unable to be pregnant. Menstruation directly impacts women’s fertility and women who see a doctor sooner for their fertility might be more likely to become pregnant in a year through assisted reproductive technology (ART) such as *in vitro* fertilization (IVF) and intrauterine insemination (IUI). We adjusted for the long-term amenorrhea when assessing for infertility because regular menstruation impacts infertility as well as blood metal concentrations. For example, the intestinal absorption of Cd, Pb, and Hg increases when the body iron stores are depleted^62^ and menstruating women are more likely to have low iron stores.^63^

### Statistical Analysis

For NHANES datasets, the use of sampling weights and sample design variables is recommended for all analyses because the sample design is both a clustered design and incorporates differential probabilities of selection. Statistical Analysis Software v9.4 (SAS Institute, Cary, NC) was used to perform all statistical analyses, incorporating sampling weights and non-responses while adjusting for cluster (PSUs) and strata of the complex sample design in NHANES.^64,65^ Weighting was calculated using NHANES sub-sample weights and were calculated according to NHANES protocols and documentation.^66^

Descriptive statistics were calculated for both outcomes and exposures: Cd, Pb, Hg, and the mixture (Mix). Statistical differences in the covariates between the outcome groups were evaluated using a chi-squared test for categorical variables and a t-test for continuous variables. Because blood concentrations of Pb, Cd, and Hg had skewed distributions based on normality tests, log transformed metal values were used. In addition to assessing the blood concentrations continuously, we also categorized the data into quartiles using the lowest quartile as the reference group. Multiple logistic regression analysis was used to evaluate the independent association between blood metal concentrations and metal mixture values and infertility after adjusting for above-mentioned covariates. The same approach was used to evaluate associations between blood metal concentrations and metal mixture values and long-term amenorrhea. Crude odds ratios (OR) and adjusted ORs and their corresponding 95% confidence intervals (CI) were presented. We used three models to examine associations between women’s blood heavy metal concentrations and historical infertility (Table 3). In model 1, crude odds ratios (OR) were calculated without adjusting for any covariates. In model 2, an adjusted model was applied by including all covariates except for the other two metals not being assessed. In model 3, a fully adjusted model was run, which included all covariates including the other two metals.

Several sensitivity analyses were conducted to examine the robustness of our findings. First, we determined that there was a difference in infertility status among the 80 additional women included in the infertility group (n=1,999) compared to the long-term amenorrhea group (n=1,919). This helped us determine that there was sufficient reason to keep both outcomes (infertility and long-term amenorrhea) as separate population groups rather than taking the smaller sample size for analysis. Second, using a chai squared test, we examined the difference in infertility status among women who may have seen a doctor and received assistance to become pregnant versus those who did not. The question of “seen a doctor because unable to become pregnant?” helped us define if an individual received medical assistance to help with her fertility or not. The purpose of this was to have additional descriptive information regarding the study population. Third, we examined the relation between long-term amenorrhea and infertility history using a chi-squared test.

## Results

### Exposure to heavy metals and women’s infertility

#### Study population

As shown in Table 1, a total of 238 or 12.8% of women were considered ever-infertile. These ever-infertile women were compared to two control groups: the main group of 1,761 women who self-reported being fertile and the sub-group of 59 pregnant women. Compared to fertile women, women who have been ever-infertile were more likely to be older, married, obese, smokers, and had seen a doctor because they were unable to become pregnant (all *p*-values < 0.05). The race/ethnicity, educational level, poverty income ratio, hormone-based contraception use, and having a period in the last 12 months were similar between ever-infertile and fertile women. Compared to pregnant women, ever-infertile women were more likely to be older, covered by health insurance, and had seen a doctor because they were unable to become pregnant (all *p*-values < 0.05). The distributions of race/ethnicity, education level, marital status, poverty income ratio, BMI, smoking, use of hormonal contraception, and having a period in the last 12 months were similar between ever-infertile and pregnant women.

**Table 1:** Women’s characteristics for studying associations between blood heavy metal concentrations and historical infertility

| Characteristics | Main group sample (ever-infertile vs fertile) |  |  |  | Sub-group sample (ever-infertile vs pregnant) |  |  |  |
| --- | --- | --- | --- | --- | --- | --- | --- | --- |
|  | Total Sample N (%) | Ever-infertile<sup>1</sup> N (%) | Fertile<sup>2</sup> N (%) | p-Value<sup>4</sup> | Total Sample N (%) | Ever-infertile<sup>1</sup> N (%) | Pregnant<sup>3</sup> N (%) | p-Value<sup>4</sup> |
| Total Subjects | 1999 | 238 (12.8) | 1761 (87.2) |  | 297 | 238 (81.6) | 59 (18.4) |  |
| Age, *mean ± SE* (years) | 34 ± 0.23 | 37 ± 0.79 | 33 ± 0.22 | <.001 | 35 ± 0.71 | 37 ± 0.70 | 27 ± 0.58 | <.001 |
| Race/Ethnicity |  |  |  | 0.85 |  |  |  | 0.26 |
| Hispanic | 518 (18.2) | 62 (17.9) | 456 (18.2) |  | 78 (18.5) | 62 (17.9) | 16 (21.4) |  |
| Non-Hispanic White | 697 (58.4) | 90 (60.5) | 607 (58.1) |  | 108 (57.8) | 90 (60.5) | 18 (45.8) |  |
| Non-Hispanic Black | 426 (13.2) | 47 (13.0) | 379 (13.2) |  | 61 (14.4) | 47 (13.0) | 14 (20.4) |  |
| Other Race Including Multi-Racial | 358 (10.2) | 39 (8.6) | 319 (10.5) |  | 50 (9.3) | 39 (8.6) | 11 (12.4) |  |
| Education Level |  |  |  | 0.53 |  |  |  | 0.73 |
| Less than High School | 297 (10.8) | 36 (11.7) | 261 (10.7) |  | 46 (15.5) | 36 (11.7) | 10 (14.1) |  |
| High School | 397 (19.6) | 50 (22.1) | 347 (19.2) |  | 62 (20.9) | 50 (22.1) | 12 (17.1) |  |
| More than High School | 1305 (69.6) | 152 (66.3) | 1153 (70.1) |  | 189 (63.6) | 152 (66.3) | 37 (68.8) |  |
| Marital Status |  |  |  | <.001 |  |  |  | 0.59 |
| Married / Living with Partner | 1173 (61.3) | 175 (78.1) | 998 (58.9) |  | 223 (78.6) | 175 (78.1) | 48 (80.6) |  |
| Divorced / Widowed / Separated | 243 (10.3) | 29 (8.8) | 214 (10.5) |  | 31 (8.0) | 29 (8.8) | 2 (4.2) |  |
| Never Married | 583 (28.4) | 34 (13.0) | 549 (30.6) |  | 43 (13.4) | 34 (13.0) | 9 (15.2) |  |
| Poverty Income Ratio, *mean ± SE* | 2.770 ± 0.07 | 2.95 ± 0.14 | 2.75 ± 0.070 | 0.15 | 2.90 ± 0.14 | 2.95 ± 0.14 | 2.72 ± 0.23 | 0.52 |
| Covered by Health Insurance |  |  |  | 0.33 |  |  |  | 0.03 |
| Yes | 1601 (83.0) | 187 (80.7) | 1414 (83.3) |  | 239 (82.6) | 187 (80.7) | 52 (91.0) |  |
| No | 398 (17.0) | 51 (19.3) | 347 (16.7) |  | 58 (17.4) | 51 (19.3) | 7 (9.0) |  |
| Body Mass Index (kg/m**2) |  |  |  | 0.007 |  |  |  | 0.16 |
| Underweight (<18.5) | 39 (2.0) | 4 (1.2) | 35 (2.1) |  | 5 (1.2) | 4 (1.2) | 1 (1.4) |  |
| Normal Weight (18.5-24.9) | 615 (32.1) | 64 (27.8) | 551 (32.7) |  | 78 (26.2) | 64 (27.8) | 14 (19.2) |  |
| Overweight (25.0-29.9) | 471 (24.9) | 38 (18.5) | 433 (25.9) |  | 53 (21.1) | 38 (18.5) | 15 (32.4) |  |
| Obesity (>30) | 874 (50.0) | 132 (52.5) | 742 (39.3) |  | 161 (51.5) | 132 (52.5) | 29 (47.0) |  |
| Ever Smoked |  |  |  | 0.028 |  |  |  | 0.50 |
| Yes | 613 (33.0) | 88 (40.5) | 525 (31.9) |  | 109 (39.3) | 88 (40.5) | 21 (34.1) |  |
| No | 1386 (67.0) | 150 (59.5) | 1236 (68.1) |  | 188 (60.7) | 150 (59.5) | 38 (65.9) |  |
| Ever taken hormone-based contraception? |  |  |  | 0.85 |  |  |  | 0.15 |
| Yes | 1389 (75.7) | 170 (76.3) | 1219 (75.6) |  | 204 (74.3) | 170 (76.3) | 34 (65.1) |  |
| No | 610 (24.3) | 68 (23.7) | 542 (24.4) |  | 93 (25.7) | 68 (23.7) | 25 (34.9) |  |
| At least one period in past 12 months |  |  |  | 0.73 |  |  |  | n/a |
| Yes | 1815 (89.8) | 222 (89.0) | 1593 (89.9) |  | 281 (91.0) | 222 (89.0) | 59 (100) |  |
| No | 183 (10.2) | 16 (11.0) | 167 (10.1) |  | 16 (9.0) | 16 (11.0) | 0 (0) |  |
| Seen a DR b/c unable to become pregnant? |  |  |  | <.001 |  |  |  | <.001 |
| Yes | 166 (9.3) | 136 (60.3) | 30 (1.8) |  | 137 (49.5) | 136 (60.3) | 1 (1.9) |  |
| No | 1833 (90.7) | 102 (39.7) | 1731 (98.2) |  | 160 (50.5) | 102 (39.7) | 58 (98.1) |  |

Values for continuous variables are mean +/- SD.
Values for categorical variables are n (unweighted sample counts) and % (weighted sample percentages to account for NHANES survey design).
<sup>1</sup> 'Ever-infertile' if subject responded 'yes' to the following question: "Have you ever attempted to become pregnant over a period of at least a year without becoming pregnant?"
<sup>2</sup> 'Fertile' if answered "No" to the following question: "Have you ever attempted to become pregnant over a period of at least a year without becoming pregnant?"
<sup>3</sup> 'Pregnant' if women answered "Yes" to the question "Are you pregnant now?"
<sup>4</sup> $p$ -Value for categorical variables comes from a chi-squared test, which determines if there is a significant difference between demographics in ever infertile vs. fertile or pregnant. $p$ -values for continuous variables comes from a t-test to determine if there is a significant difference between the means of ever infertile vs. fertile or pregnant.

The question “seen a DR b/c unable to become pregnant?” enabled us to define if a woman received medical assistance to achieve pregnancy. In the main group, 166 (8.3%) women reported seeing a doctor of which 136 (6.8%) were ever-infertile compared to 30 (1.5%) who self-reported to be fertile. In the sub-group, 137 (46.1%) women reported seeing a doctor of which 136 (45.8%) were ever-infertile compared to only one woman (0.3%) who was pregnant. Women who had seen a doctor were substantially more likely to be ever-infertile in both the main group and sub-group women (*p*-value <.001).

#### Bivariate results and metal exposures

The median and log transformed means of blood heavy metal concentrations in women with various fertility status are summarized in Table 2 and illustrated in Figures 3 and 4. With respect to the main group analysis, there was no significant difference for the blood concentrations of all three single heavy metals and mixtures between ever-infertile and self-reported fertile women (Figure 3). In the sub-group analysis, women who have been ever-infertile had significantly higher concentrations of blood Pb and heavy metal mixture than pregnant women (Table 2 and Figure 4). The blood concentrations of Cd and Hg, however, were comparable in the main and sub-groups (Table 2 and Figure 3 and 4).

**Table 2:**
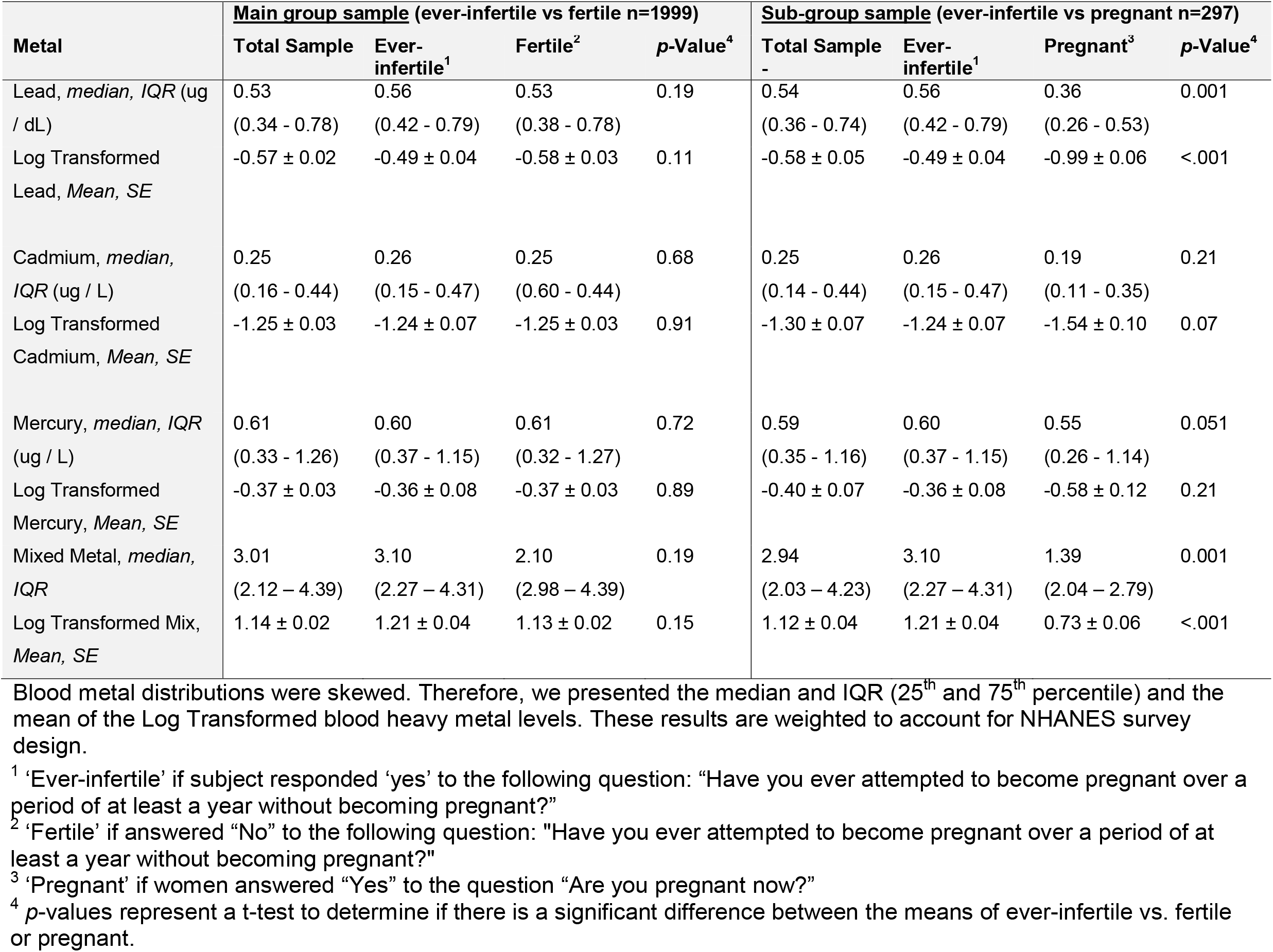
Medians and log transformed means of blood heavy metal concentrations and heavy metal mixture scores in ever-infertile or fertile/pregnant women

**Figure 3:**
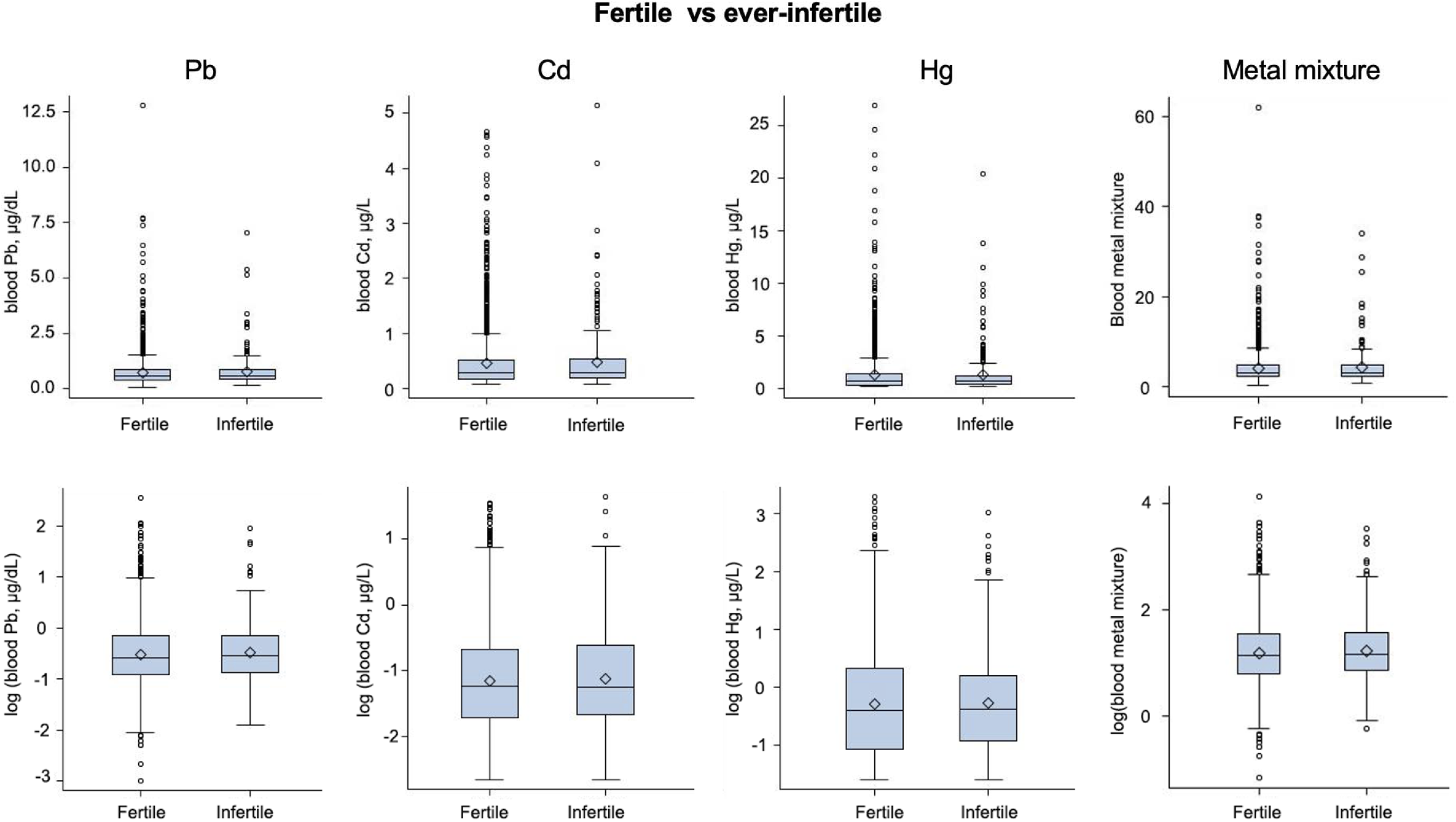
The original and log-transformed blood heavy metal concentrations and heavy metal mixture scores in women for the main-group (ever-infertile and fertile) comparison. Each box plot includes the lower (25%) and upper (75%) quartile, median (string), and mean (diamond dot). These results are un-weighted.

**Figure 4:**
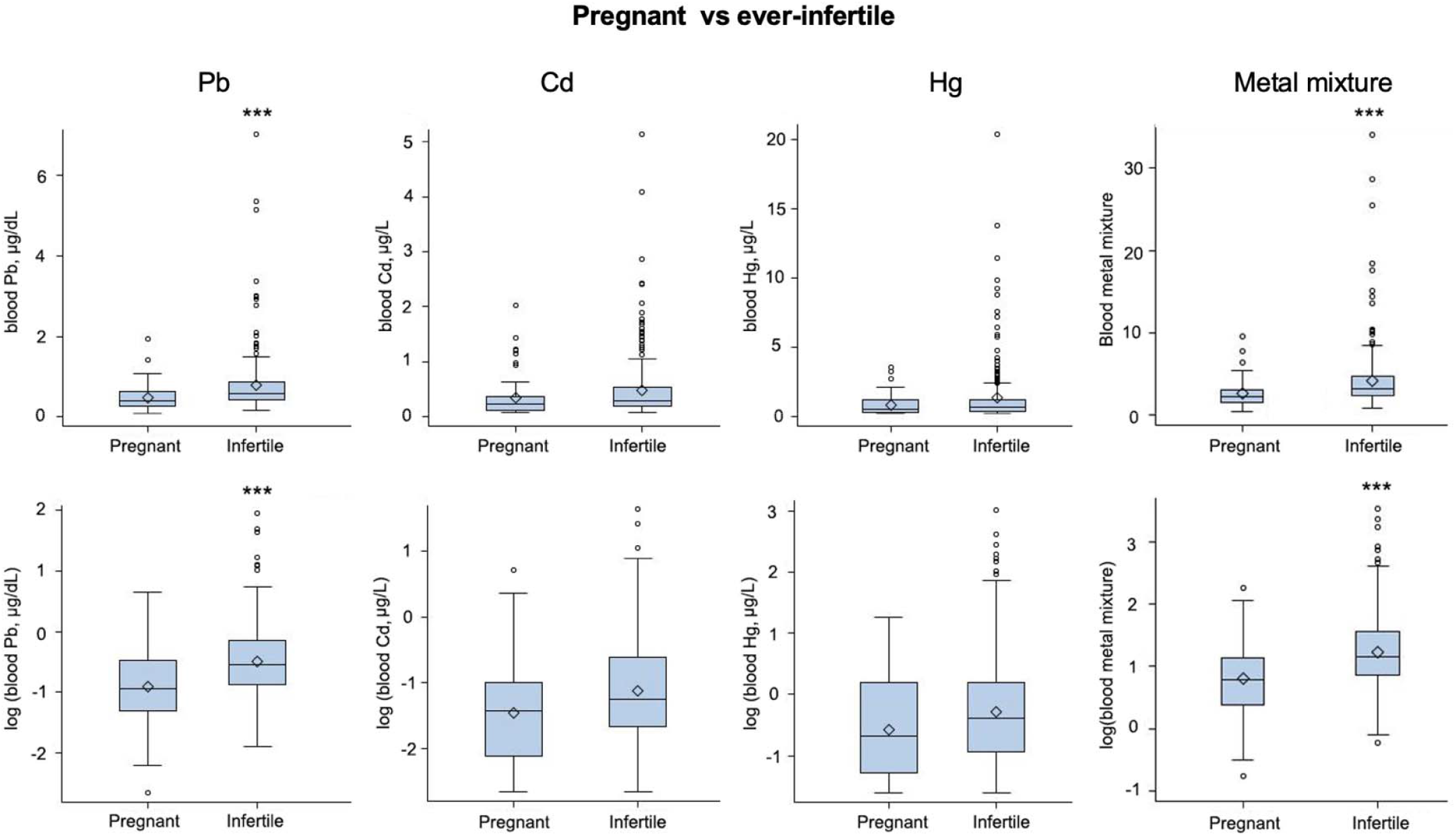
The blood heavy metal distributions among the sub-group (pregnant and ever-infertile) samples. These results are un-weighted. The original and log-transformed blood heavy metal concentrations and heavy metal mixture scores in women for the sub-group (pregnant and fertile) comparison. Each box plot includes the lower (25%) and upper (75%) quartile, median (string), and mean (diamond dot). These results are un-weighted. \*\*\**p*<0.001.

#### Multiple logistic regression analysis results

Multiple logistic regression analysis showed that after full adjustment including demographic characteristics, lifestyle factors, and two metals not being assessed (model 3), there was a positive association between blood Pb concentrations and women’s ever-infertility. The continuous log transformed data of both the main-group and sub-group analyses showed that as blood Pb concentrations increased, women were more likely to be ever-infertile (OR: 1.75, 95% CI: 1.01-3.02; and OR: 3.09, 95% CI: 1.22-7.85, respectively, Table 3). The results of model 1 with crude OR and model 2 with adjustments of all covariates but not two metals not being assessed showed similar results, except that the crude OR of the main group analysis is insignificant (Table 3).

**Table 3:** Associations between blood heavy metal concentrations and heavy metal mixture scores and women’s infertility

| Characteristics | Total<br>N (%)<sup>1</sup> | Main group sample (ever infertile<sup>2</sup> vs fertile<sup>3</sup><br>n=1999) |  |  |  | Sub-group sample (ever-infertile<sup>2</sup> vs pregnant<sup>4</sup><br>n=297) |  |  |  |
| --- | --- | --- | --- | --- | --- | --- | --- | --- | --- |
|  |  | Ever-infertile<sup>2</sup><br>n (%) or<br>Mean (SD)<sup>1</sup> | Crude OR<br>(95% CI)<br>Model 1 | Adj OR<br>(95% CI)<br>Model 2 | Fully Adj<br>OR (95%<br>CI)<br>Model 3 | Ever-infertile<sup>2</sup><br>n (%) or<br>Mean (SD)<sup>1</sup> | Crude OR<br>(95% CI)<br>Model 1 | Adj OR<br>(95% CI)<br>Model 2 | Fully Adj<br>OR (95%<br>CI)<br>Model 3 |
| LEAD |  |  |  |  |  |  |  |  |  |
| Log Transformed,<br>Mean (SD) | -0.52<br>(0.62) | -0.48 (0.61) | 1.28 (0.97<br>- 1.68) | 1.69<br>(1.01 -<br>2.85)* | 1.75 (1.02 -<br>3.02)* | -0.48 (0.61) | 5.02 (2.73<br>- 9.23)* | 5.19 (2.14 -<br>12.59)* | 3.09 (1.22 -<br>7.85)* |
| Lead quartiles, n<br>(%) |  |  |  |  |  |  |  |  |  |
| Q1 (Ref) (≤0.40) | 488<br>(24.41%) | 52 (2.6%) | Ref | Ref | Ref | 52 (17.51%) | Ref | Ref | Ref |
| Q2 (0.41 – 0.56) | 492<br>(24.61%) | 56 (2.8%) | 1.15 (0.70<br>- 1.89) | 1.37<br>(0.67 -<br>2.80) | 1.38 (0.68 -<br>2.79) | 56 (18.86%) | 3.23 (1.18<br>- 8.86)* | 2.88 (0.61 -<br>13.55) | 2.52 (0.53 -<br>12.09) |
| Q3 (0.57 – 0.86) | 513<br>(25.66%) | 70 (3.5%) | 1.52 (1.00<br>- 2.29)* | 1.60<br>(0.82 -<br>3.10) | 1.61 (0.83 -<br>3.09) | 70 (23.57%) | 5.32 (2.20<br>- 12.88)* | 5.60 (1.67 -<br>18.73)* | 3.47 (1.11 -<br>10.83)* |
| Q4 (>0.86) | 506<br>(25.31) | 60 (3.0%) | 1.30 (0.80<br>- 2.11) | 1.71<br>(0.76 -<br>3.85) | 1.72 (0.75 -<br>3.95) | 60 (20.2%) | 6.71 (2.85<br>- 15.81)* | 12.62 (2.48<br>- 64.21)* | 5.26 (1.18 -<br>23.54)* |
| CADMIUM |  |  |  |  |  |  |  |  |  |
| Log Transformed,<br>Mean (SD) | -1.16<br>(0.84) | -1.13 (0.84) | 1.01 (0.81<br>- 1.26) | 1.01<br>(0.68 -<br>1.52) | 0.94 (0.63 -<br>1.41) | -1.13 (0.84) | 1.51 (0.94<br>- 2.43) | 2.29 (0.87 -<br>6.04) | 1.90 (0.64 -<br>5.63) |
| Cadmium quartiles,<br>n (%) |  |  |  |  |  |  |  |  |  |
| Q1 (Ref) ($\leq 0.18$) | 469<br>(23.46%) | 52 (2.6%) | Ref | Ref | Ref | 52 (17.51%) | Ref | Ref | Ref |
| Q2 (0.19 – 0.29) | 509<br>(25.46%) | 67 (3.35%) | 1.08 (0.66<br>- 1.77) | 0.65<br>(0.34 -<br>1.25) | 0.67 (0.35 -<br>1.27) | 67 (22.56%) | 2.06 (0.81<br>- 5.27) | 0.82 (0.16 -<br>4.05) | 0.58 (0.10 -<br>3.41) |
| Q3 (0.30-0.51) | 513<br>(25.66%) | 53 (2.65%) | 0.87 (0.57<br>- 1.32) | 0.71<br>(0.38 -<br>1.33) | 0.70 (0.37 -<br>1.31) | 53 (17.85%) | 1.83 (0.76<br>- 4.41) | 0.38 (0.05 -<br>2.83) | 0.37 (0.04 -<br>3.20) |
| Q4 ($>0.51$) | 508<br>(25.41%) | 66 (3.3%) | 1.07 (0.64<br>- 1.81) | 0.95<br>(0.41 -<br>2.18) | 0.83 (0.36 -<br>1.94) | 66 (22.22%) | 2.19 (0.80<br>- 6.02) | 1.33 (0.09 -<br>19.70) | 0.61 (0.04 -<br>9.94) |
| **MERCURY** |  |  |  |  |  |  |  |  |  |
| Log Transformed,<br>Mean (SD) | -0.29<br>(0.99) | -0.28 (0.96) | 1.01 (0.85<br>- 1.21) | 1.07<br>(0.80 -<br>1.44) | 1.02 (0.76 -<br>1.36) | -0.28 (0.96) | 1.35 (0.84<br>- 2.17) | 1.37 (0.71 -<br>2.67) | 1.38 (0.76 -<br>2.51) |
| Mercury quartiles, n<br>(%) |  |  |  |  |  |  |  |  |  |
| Q1 (Ref) ($\leq 0.34$) | 491<br>(24.56%) | 46 (2.3%) | Ref | Ref | Ref | 46 (15.49%) | Ref | Ref | Ref |
| Q2 (0.34 – 0.67) | 508<br>(25.41%) | 72 (3.6%) | 1.69 (0.96<br>- 2.97) | 1.70<br>(0.76 -<br>3.81) | 1.59 (0.70 -<br>3.61) | 72 (24.24%) | 2.54 (0.90<br>- 7.17) | 2.26 (0.47 -<br>10.90) | 1.48 (0.43 -<br>5.03) |
| Q3 (0.68-1.38) | 493<br>(24.66%) | 63 (3.15%) | 1.43 (0.82<br>- 2.49) | 1.54<br>(0.75 -<br>3.18) | 1.49 (0.72 -<br>3.08) | 63 (21.21%) | 1.91 (0.62<br>- 5.90) | 2.53 (0.54 -<br>11.78)* | 2.51 (0.60 -<br>10.59) |
| Q4 (>1.38) | 507<br>(25.36%) | 57 (2.85%) | 1.20 (0.66<br>- 2.17) | 0.58<br>(3.66 -<br>3.11) | 1.26 (0.51 -<br>3.11) | 57 (19.19%) | 1.90 (0.61<br>- 5.94) | 2.01 (0.40 -<br>10.20) | 1.67 (0.33 -<br>8.43) |
| **MIX** |  |  |  |  |  |  |  |  |  |
| Log Transformed,<br>Mean (SD) | 1.19<br>(0.60) | 1.22 (0.61) | 1.26 (0.94<br>- 1.69) | 1.69<br>(0.95 -<br>2.99) | 1.00 (0.46 -<br>2.19) | 1.22 (0.61) | 4.60 (2.42<br>- 8.76)* | 6.21 (2.24 -<br>17.20) * | 1.30 (0.09 -<br>18.13) |
| Mix quartiles, n (%) |  |  |  |  |  |  |  |  |  |
| Q1 (Ref) ($\leq 2.24$) | 499<br>(24.96%) | 53 (2.65%) | Ref | Ref | Ref | 53 (17.85%) | Ref | Ref | Ref |
| Q2 (2.24-3.14) | 500<br>(25.01%) | 65 (3.25%) | 1.34 (0.91<br>- 1.99) | 1.58<br>(0.86 -<br>2.91) | 1.47 (0.80 -<br>2.71) | 65 (21.89%) | 2.66 (1.33<br>- 5.32) * | 1.86 (0.53 -<br>6.47) | 0.87 (0.18 -<br>4.30) |
| Q3 (3.14-4.74) | 500<br>(25.01%) | 59 (2.95%) | 1.39 (0.90<br>- 2.15) | 1.31<br>(0.65 -<br>2.63) | 1.12 (0.56 -<br>2.25) | 59 (19.87%) | 9.62 (2.74<br>- 33.80) * | 15.87 (3.11<br>- 80.91) * | 6.92 (0.72 -<br>66.79) |
| Q4 (>4.74) | 500<br>(25.01%) | 61 (3.05%) | 1.29 (0.82<br>- 2.05) | 2.02<br>(0.89 -<br>4.75) | 1.17 (0.44 -<br>3.10) | 61 (20.54%) | 4.80 (1.77<br>- 13.03) * | 13.22 (2.39<br>- 73.17) * | 0.48 (0.01 -<br>16.32) |

\* Statistically significant and corresponding $p$ -value $<0.05$
<sup>1</sup> Values are unweighted sample counts and percentages.
<sup>2</sup> 'Ever infertile' if subject responded 'yes' to the following question: "Have you ever attempted to become pregnant over a period of at least a year without becoming pregnant?"
<sup>3</sup> 'Fertile' if answered "No" to the following question: "Have you ever attempted to become pregnant over a period of at least a year without becoming pregnant?"
<sup>4</sup> 'Pregnant' if women answered "Yes" to the question "Are you pregnant now?"

Multiple logistic regression results for the categorical data in model 3 revealed that there was no association between Pb and infertility for all quartiles of 2, 3 and 4 compared with the lowest quartile 1 in the main group analysis (ever-infertile vs. fertile). However, for the sub-group analysis (ever-infertile vs. pregnant), the blood concentrations of Pb in quartiles 3 and 4 were significantly associated with women’s historical infertility (OR: 3.47, 95% CI: 1.11-10.83; and OR: 5.26, 95% CI: 1.18-23.54, respectively), and the OR from quartiles 2 to 4 exhibited a dose-dependent relationship (Table 3). The results of model 1 with crude OR and model 2 with adjustments of all covariates but not two metals not being assessed showed similar results (Table 3).

With respect to Cd and Hg summarized in Table 3, the results of both continuous and categorical multiple logistic regression analyses in all three models revealed no significant associations except that the increase of blood concentrations of Hg in the quartile 3 was significantly associated with women’s infertility in model 2 of the sub-group analysis (OR: 2.53, 95% CI: 0.64-11.78). Regarding the heavy metal mixture, model 3 showed no significant associations between the metal mixture and women’s infertility in both the main and sub-group analyses. Contrarily, sub-group analysis in models 1 and 2 revealed that metal mixtures are positively associated with women’s ever-infertility; since models 1 and 2 did not adjust for the single metals, this is likely due to the positive association found between Pb and infertility. Collectively, these results indicate that after full adjustment, exposure to Pb increases the odds of women’s historical infertility; further, no associations were found between Cd, Hg, and the mixture of all three metals and women’s historical infertility.

### Women’s historical infertility is not associated with their recent long-term amenorrhea

We next examined associations between women’s historical infertility and recent long-term amenorrhea. A total of 1,918 women had complete data of both infertility and long-term amenorrhea (Table 4). There was no statistical correlation between women’s long-term amenorrhea and historical infertility (p-value = 0.29), although the percentage of long-term amenorrhea in women who were ever-infertile (3.9%) was slightly lower than that in fertile women (5.6%) and the percentage of historical infertility in women with long-term amenorrhea (8.7%) was lower than that in menstruating women (12.2%). This negative association suggests that women’s historical infertility does not reflect their recent reproductive status. The NHANES survey asked “*Have you had at least one menstrual period in the past 12 months?*” Because the absence of a period or amenorrhea for 12 consecutive months has been suggested as an important indicator of menopause,^57^ the long-term amenorrhea may reflect women’s most recent reproductive and fertility status. Thus, as a secondary outcome, we chose to investigate associations between heavy metal exposure and women’s recent long-term amenorrhea.

**Table 4:** Association between women’s infertility and recent long-term amenorrhea

|  | **Amenorrhea** | **Menstruating** | **Total** | **% of women with amenorrhea by infertility status** | **p-value** |
| --- | --- | --- | --- | --- | --- |
| **Infertile** | 9 | 222 | 231 | 3.9% | 0.29 |
| **Fertile** | 94 | 1593 | 1687 | 5.6% |  |
| **Total** | 103 | 1815 | 1918 |  |  |
| **% of women who are infertile by menstrual status** | 8.7% | 12.2% |  |  |  |

### Exposure to heavy metals and women’s long-term amenorrhea

#### Study population

As shown in Figure 2, a total of 1,919 participants were included to assess long-term amenorrhea after further deleting participants with missing data on long-term amenorrhea (n=361), demographic variables (n=190), BMI (n=12), and information on the use of birth control pill and female hormone use (n=4). The characteristics of these women are summarized in Table 5. Compared with menstruating women, women with long-term amenorrhea were more likely to be Non-Hispanic White and Non-Hispanic Black (*p*-value < 0.05) compared to other ethnicities. However, the distributions of age, educational level, marital status, health insurance coverage, poverty income ratio, BMI, smoking history, and hormone-based contraception use were largely similar between menstruating women and women with long-term amenorrhea (all *p-*values > 0.05).

**Table 5:** Women’s characteristics for studying associations between blood heavy metal concentrations and long-term amenorrhea

| Characteristics | Total Sample<br>N (%) | Long-term<br>amenorrhea<sup>1</sup><br>N (%) | Menstruating<sup>2</sup><br>N (%) | p-Value<sup>3</sup> |
| --- | --- | --- | --- | --- |
| Total Subjects | 1919 | 103 (6.4) | 1816 (93.6) |  |
| Age, *mean ± SE* (years) | 34 ± 0.2 | 35 ± 0.1 | 33 ± 0.2 | 0.24 |
| Race/Ethnicity |  |  |  | 0.03 |
| Hispanic | 501 (18.4) | 23 (11.9) | 478 (18.8) |  |
| Non-Hispanic White | 664 (58.0) | 47 (68.5) | 617 (57.3) |  |
| Non-Hispanic Black | 409 (13.4) | 26 (14.1) | 383 (13.3) |  |
| Other Race Including Multi-Racial | 345 (10.3) | 7 (5.6) | 338 (10.6) |  |
| Education Level |  |  |  | 0.65 |
| Less than High School | 285 (10.7) | 16 (11.8) | 269 (10.6) |  |
| High School | 382 (19.9) | 27 (23.0) | 355 (19.6) |  |
| More than High School | 1252 (69.4) | 60 (65.3) | 1192 (69.7) |  |
| Marital Status |  |  |  | 0.77 |
| Married / Living with Partner | 1117 (60.8) | 57 (63.5) | 1060 (60.6) |  |
| Divorced / Widowed / Separated | 228 (10.1) | 13 (10.8) | 215 (10.1) |  |
| Never Married | 574 (29.1) | 33 (25.7) | 541 (29.3) |  |
| Covered by Health Insurance |  |  |  | 0.47 |
| Yes | 1533 (82.5) | 88 (85.7) | 1445 (82.3) |  |
| No | 386 (17.5) | 15 (14.3) | 371 (17.7) |  |
| Poverty Income Ratio, *mean ± SE* | 2.75 ± 0.065 | 2.59 ± 0.189 | 2.77 ± 0.067 | 0.39 |
| Body Mass Index (kg/m**2) |  |  |  | 0.88 |
| Underweight (<18.5) | 38 (2.1) | 1 (1.2) | 37 (2.1) |  |
| Normal Weight (18.5-24.9) | 591 (31.8) | 34 (31.9) | 557 (31.8) |  |
| Overweight (25.0-29.9) | 451 (25.0) | 21 (22.6) | 430 (25.2) |  |
| Obesity (>30) | 839 (41.1) | 47 (44.4) | 792 (40.9) |  |
| Ever Smoked |  |  |  | 0.51 |
| Yes | 580 (32.6) | 34 (35.9) | 546 (32.3) |  |
| No | 1339 (67.4) | 69 (64.1) | 1270 (67.7) |  |
| Ever taken hormone-based contraception? |  |  |  | 0.38 |
| Yes | 1328 (75.4) | 73 (71.3) | 1255 (75.7) |  |
| No | 591 (24.6) | 30 (28.7) | 561 (24.3) |  |

Values for continuous variables are mean +/- the Standard Error of the Mean.
Values for categorical variables are n (unweighted sample counts) and % (weighted sample percentages to account for NHANES survey design).
If precents do not equal 100% it is due to rounding.
<sup>1</sup> 'Long-term amenorrhea' answered "no" to the question "Have you had at least one menstrual period in the past 12 months? (Please do not include bleedings caused by medical conditions, hormone therapy, or surgeries.)" AND answered "Other" or "Don't know" to the question "What is the reason that you have not had a period in the past 12 months?"
<sup>2</sup> 'Menstruating' if answered "Yes" to the question "Have you had at least one menstrual period in the past 12 months? (Please do not include bleedings caused by medical conditions, hormone therapy, or surgeries.)"
<sup>3</sup> P-Value for categorical variables comes from a chi-squared test, which determines if there is a significant difference between demographics in long-term amenorrhea vs. menstruating women. P-values for continuous variables comes from a t test to determine if there is a significant difference between the means of long-term amenorrhea vs. menstruating.

#### Bivariate results and heavy metal exposures

The median and log transformed means of blood heavy metal concentrations are shown in Table 6 and illustrated in Figure 5. Compared with menstruating women, women with long-term amenorrhea had comparable blood concentrations of Pb, Cd, and heavy metal mixtures but had significantly higher median blood concentrations of Hg (Figure 5).

**Table 6:** Medians and log transformed means of blood heavy metal concentrations and heavy metal mixture scores in women with normal menstruation and long-term amenorrhea

| **Metal** | **Total Sample** | **Long-term amenorrhea<sup>1</sup>** | **Menstruating<sup>2</sup>** | **p-Value<sup>3</sup>** |
| --- | --- | --- | --- | --- |
| Lead, *median, IQR* (ug / dL) | 0.53 (0.38 - 0.78) | 0.52 (0.35 - 0.71) | 0.53 (0.38 - 0.78) | 0.72 |
| Log Transformed Lead, *Mean, SE* | -0.58 ± 0.02 | -0.62 ± 0.09 | -0.57 ± 0.02 | 0.60 |
| Cadmium, *median, IQR* (ug / L) | 0.25 (0.16 - 0.44) | 0.23 (0.12 - 0.41) | 0.26 (0.16 - 0.44) | 0.99 |
| Log Transformed Cadmium, *Mean, SE* | -1.25 ± 0.03 | -1.37 ± 0.11 | -1.25 ± 0.03 | 0.33 |
| Mercury, *median, IQR* (ug / L) | 0.61 (0.33 - 1.26) | 0.65 (0.37 - 1.01) | 0.61 (0.32 - 1.29) | 0.004 |
| Log Transformed Mercury, *Mean, SE* | -0.37 ± 0.03 | -0.38 ± 0.06 | -0.37 ± 0.03 | 0.84 |
| Mixed Metal, *median, IQR* | 2.97 (2.10 – 4.34) | 2.91 (1.84 – 3.82) | 2.97 (2.10 – 4.37) | 0.74 |
| Log Transformed Mix, *Mean, SE* | 1.13 ± 0.02 | 1.08 ± 0.08 | 1.13 ± 0.02 | 0.57 |

Blood metal distributions were skewed. Therefore, we presented the median and IQR (25<sup>th</sup> and 75<sup>th</sup> percentile) and the mean of the Log Transformed blood metal levels. These results are weighted to account for NHANES survey design.
<sup>1</sup> ‘Long-term amenorrhea’ answered “no” to the question “Have you had at least one menstrual period in the past 12 months? (Please do not include bleedings caused by medical conditions, hormone therapy, or surgeries.)” AND answered “Other” or “Don’t know” to the question “What is the reason that you have not had a period in the past 12 months?”
<sup>2</sup> ‘Menstruating’ if answered “Yes” to the question “Have you had at least one menstrual period in the past 12 months? (Please do not include bleedings caused by medical conditions, hormone therapy, or surgeries.)”
<sup>3</sup> P-Values represent a t test to determine if there is a significant difference between the means of long-term amenorrhea vs. menstruating.

**Figure 5:**
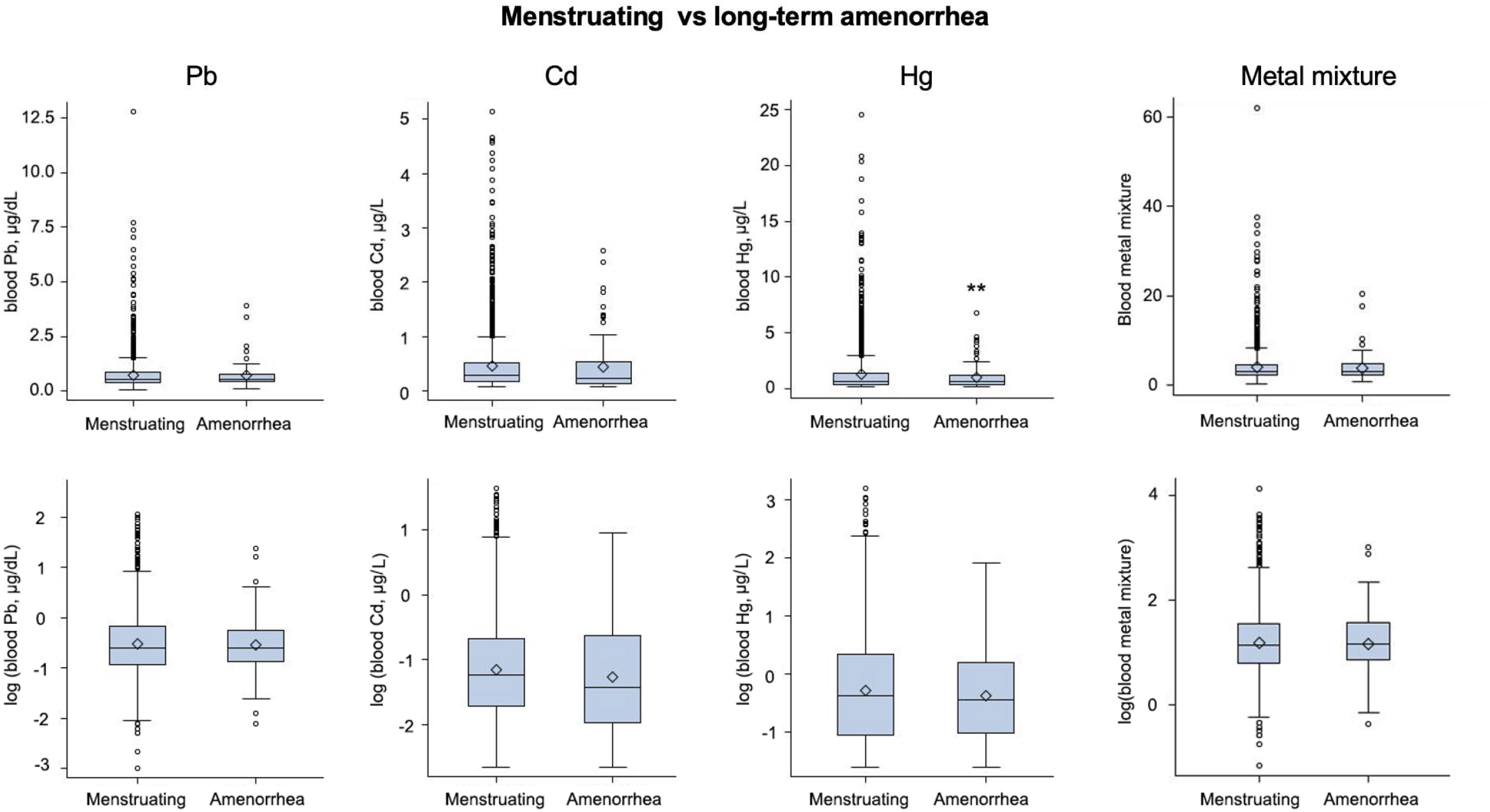
The original and log-transformed blood heavy metal concentrations and heavy metal mixture scores in women with normal menstruation and long-term amenorrhea. Each box plot includes the lower (25%) and upper (75%) quartile, median (string), and mean (diamond dot). These results are un-weighted.

#### Multiple logistic regression model results

Table 7 summarizes associations between blood heavy metal concentrations and women’s long-term amenorrhea assessed by three models as we described in the fertility analysis. Multiple logistic regression analysis from continuous and categorical data showed no significant associations between blood concentrations of Pb or Hg and women’s long-term amenorrhea in all three models. In the categorical multiple logistic regression analysis, after the full adjustment in model 3, there was a negative association between the blood Cd concentrations in quartiles 2 and 3 and women’s long-term amenorrhea (quartile 2 OR: 0.47, 95% CI: 0.25-0.87; quartile 3 OR: 0.31, 95% CI: 0.13-0.76). Similar to model 3, the results of model 1 and 2 also showed an inverse association between the blood concentrations of Cd in quartiles 2 and 3 and long-term amenorrhea. The ORs, although still less than 1, were higher for quartile 4 than those for quartiles 2 and 3 in all three models but were not statically significant (Table 7). For the mixture of all three heavy metals, all three models showed insignificant associations between blood metal mixture concentrations and long-term amenorrhea.

**Table 7:** Associations between blood heavy metal concentrations and heavy metal mixture scores and women’s long-term amenorrhea

| Characteristics | Total N (%)<sup>1</sup> | Full sample (Long-term amenorrhea<sup>1</sup> vs Menstruating<sup>2</sup> n=1919) |  |  |  |
| --- | --- | --- | --- | --- | --- |
|  |  | Long-term amenorrhea n (%) or Mean (SD) | Crude OR (95% CI) Model 1 | Adj OR (95% CI) Model 2 | Fully Adj OR (95% CI) Model 3 |
| LEAD |  |  |  |  |  |
| Log Transformed, Mean (SD) | -0.53 (0.62) | -0.54 (0.56) | 0.88 (0.53 - 1.46) | 0.89 (0.53 - 1.50) | 0.93 (0.54 - 1.59) |
| Quartiles, n (%) |  |  |  |  |  |
| Q1 (≤0.39) | 460 (23.97%) | 22 (1.15%) | Ref | Ref | Ref |
| Q2 (0.40 - 0.55) | 475 (24.75%) | 27 (1.41%) | 0.99 (0.41 - 2.35) | 1.02 (0.42 - 2.50) | 1.04 (0.42 - 2.54) |
| Q3 (0.56 - 0.83) | 504 (26.26%) | 32 (1.67%) | 1.03 (0.46 - 2.32) | 1.03 (0.46 - 2.32) | 1.06 (0.46 - 2.42) |
| Q4 (>0.83) | 480 (25.01%) | 22 (1.15%) | 0.71 (0.31 - 1.61) | 0.72 (0.31 - 1.68) | 0.76 (0.31 - 1.85) |
| CADMIUM |  |  |  |  |  |
| Log Transformed, Mean (SD) | -1.16 (0.83) | -1.27 (0.94) | 0.84 (0.58 - 1.22) | 0.73 (0.48 - 1.09) | 0.72 (0.49 - 1.08) |
| Quartiles, n (%) |  |  |  |  |  |
| Q1 (≤0.18) | 455 (23.71%) | 38 (1.98%) | Ref | Ref | Ref |
| Q2 (0.18 - 0.28) | 490 (25.53%) | 23 (1.20%) | 0.49 (0.26 - 0.91)* | 0.47 (0.25 - 0.87)* | 0.47 (0.25 - 0.87)* |
| Q3 (0.29 - 0.51) | 488 (25.43%) | 16 (0.83%) | 0.36 (0.16 - 0.82)* | 0.31 (0.13 - 0.75)* | 0.31 (0.13 - 0.76)* |
| Q4 (>0.51) | 486 (25.33%) | 26 (1.35%) | 0.69 (0.34 - 1.38) | 0.52 (0.23 - 1.20) | 0.53 (0.24 - 1.21) |
| MERCURY |  |  |  |  |  |
| Log Transformed, Mean (SD) | -0.29 (0.98) | -0.38 (-1.61) | 0.98 (0.84 - 1.15) | 1.10 (0.92 - 1.32) | 1.11 (0.91 - 1.36) |
| Quartiles, n (%) |  |  |  |  |  |
| Q1 ($\leq 0.34$) | 471 (24.54%) | 25 (1.30%) | Ref | Ref | Ref |
| Q2 (0.35 – 0.68) | 485 (25.27%) | 30 (1.56%) | 1.60 (0.82 - 3.13) | 1.68 (0.83 - 3.42) | 1.70 (0.84 - 3.44) |
| Q3 (0.69 – 1.39) | 480 (25.01%) | 27 (1.41%) | 1.60 (0.81 - 3.19) | 1.87 (0.92 - 3.82) | 1.87 (0.92 - 3.77) |
| Q4 ($> 1.39$) | 483 (25.17%) | 21 (1.09%) | 0.91 (0.53 - 1.56) | 1.18 (0.65 - 2.14) | 1.19 (0.63 - 2.25) |
| **MIX** |  |  |  |  |  |
| Log Transformed,<br>Mean (SD) | 1.18 (0.60) | 1.17 (0.56) | 0.86 (0.51 - 1.46) | 0.86 (0.50 - 1.47) | 0.82 (0.35 - 1.93) |
| Quartiles, n (%) |  |  |  |  |  |
| Q1 ($\leq 2.21$) | 479 (24.96%) | 24 (1.25%) | Ref | Ref | Ref |
| Q2 (2.21 – 3.10) | 480 (25.01%) | 25 (1.30%) | 0.71 (0.31 - 1.63) | 0.71 (0.30 - 1.67) | 0.74 (0.29 - 1.85) |
| Q3 (3.10 – 4.66) | 480 (25.01%) | 27 (1.41%) | 0.86 (0.39 – 1.90) | 0.86 (0.39 - 1.92) | 0.93 (0.37 - 2.34) |
| Q4 ($> 4.66$) | 480 (25.01%) | 27 (1.41%) | 0.83 (0.40 - 1.74) | 0.85 (0.39 - 1.84) | 1.04 (0.42 - 2.58) |

\*Statistically significant and corresponding $p$ -value $< 0.05$
Values are unweighted sample counts and percentages.
<sup>1</sup> 'Long-term amenorrhea' answered "no" to the question "Have you had at least one menstrual period in the past 12 months? (Please do not include bleedings caused by medical conditions, hormone therapy, or surgeries.)" AND answered "Other" or "Don't know" to the question "What is the reason that you have not had a period in the past 12 months?"
<sup>2</sup> 'Menstruating' if answered "Yes" to the question "Have you had at least one menstrual period in the past 12 months? (Please do not include bleedings caused by medical conditions, hormone therapy, or surgeries.)"

## Discussion

About 10-15% of women of reproductive age experience infertility.^67,68^ Accumulating evidence reveals the endocrine disrupting effects of heavy metals, suggesting their possible contributions to women’s impaired fertility and other reproductive disorders. Here, we performed a cross-sectional analysis of NHANES 2013-2018 to investigate associations between exposure to single Cd, Pb, Hg and mixtures and women’s infertility and long-term amenorrhea. Our results show that (1) the blood concentrations of Pb and heavy metal mixtures were significantly higher in ever-infertile women than pregnant women, but the concentrations of Cd and Hg were comparable; (2) exposure to Pb is positively associated with women’s historical infertility; and (3) the increase of blood concentrations of Cd is inversely related to women’s recent long-term amenorrhea.

### Comparisons of blood heavy metal levels between this study and guidelines from federal or other organizations

So far, there are no recognized biological functions of Pb, Cd, and Hg for human health. The typical blood levels of Pb in adults is less than 1 μg/dL, and 5 μg/dL is designated as the elevated blood lead level in adults by the US CDC.^69^ This is also the level for required medical removal in the workplace if occupational exposures exist for women who are pregnant or are trying to be pregnant due possible reproductive and developmental adversities.^70^ In our study, 81.5% of women had blood Pb levels < 1 μg/dL, 17.9% had levels at 1-5 μg/dL, and 11 women (0.55%) had levels > 5 μg/dL. The blood levels of Cd are usually < 5 μg/L, with most in the range of 0.5-2 μg/L; Blood Cd levels of 50 μg/L or more have been shown to cause acute toxicities.^71,72^ The women’s blood concentrations of Cd in our study ranged from 0.07 - 5.14 μg/L, with only one woman having blood Cd levels > 5 μg/L and 97.2% had levels < 2 μg/L. The blood concentrations of Hg are usually < 10 μg/L. Significant exposure is defined when the concentration is > 50 μg/L if exposure is due to alkyl Hg, or > 200 μg/L if exposure is due to Hg(2+).^73^ In our study, women’s blood Hg concentrations ranged from 0.2 - 26.87 μg/L, with 99.1% of them having Hg levels <10 μg/L and 18 women (0.9%) having blood Hg levels >10 μg/L. Altogether, the percentages of women that exceeded typical or normal levels of blood heavy metals were 18.5% for Pb, 0.05% for Cd, and 0.9% for Hg. Observed elevated blood heavy metal levels, particularly for Pb, pose a threat to women’s reproductive health and fertility, highlighting an urgent unmet need to prevent and reduce heavy metal exposure.

### Impacts of heavy metal exposure and women’s fertility and menstrual cycle

The impacts of heavy metal exposure on women’s fertility and menstrual outcomes remain elusive. Consistent to our data, a cross-sectional study in Taiwan from Lei et al. and another cross-sectional analysis by Lee et al. using NHANES 2013-2016 found that the blood concentrations of Pb in ever-infertile women were significantly higher than pregnant woman and this association was dose-dependent.^26,27^ Similar to Lee et al., we also found a positive association between the log transformed Pb concentrations and women’s infertility, but we found a negative association between Cd and long-term amenorrhea after quarterization. We also discovered similar results to another NHANES 2013-2016 analysis that found no associations between Hg and infertility.^74^

The absorption, distribution, metabolism, and excretion (ADME) of metals, particularly Cd, depend on nutritional status. The intestinal absorption of Cd increases when the body iron stores are depleted.^62^ In addition, women typically have higher levels of Cd than men because women are more susceptible to having low iron stores due to the monthly menstruation.^63,75^ It has also been found that people with vegan/vegetarian diets often have low iron, while concurrently these people on vegan/vegetarian diets tend to have higher blood levels of Cd.^76^ These results suggest that although we did not anticipate Cd being protective against women’s long-term amenorrhea, it is possible that women who have normal menstruation and thus the metal transporters in the GI track are more upregulated than amenorrhea women tend to have higher blood levels of Cd, resulting in a negative association in our analysis. Therefore, future research is necessary to consider associations between Cd levels, dietary patterns, iron levels, and amenorrhea.

So far, evidence regarding the effects of heavy metal exposure on women’s reproduction is limited and inconsistent; however, the rationale behind our observed associations can be explained by previous *in vitro* and *in vivo* studies.^23,77-81^ With respect to Pb, results from experimental research suggest that Pb may impact female fertility through various mechanisms, including disrupting menstrual cycle, altering hormone levels, and impairing fetal development.^82,83^ It was also found in mice that Pb accumulates in the ovary and disrupts folliculogenesis, decreases ovarian reserve, and increases follicle atresia,^79,81,84,85^ suggesting that all these Pb-induced reproductive toxicities may contribute to women’s historical infertility observed in our NHANES analysis.

Animal studies found that Cd may adversely impact female reproduction.^85^ For example, Cd has been shown to decrease the number of growing follicles,^85-87^ induce follicle atresia,^85,88^ alter follicular cell structure,^85,89,90^ decrease ovarian reserve,^85,91,92^ reduce FSH and LH levels,^85,93^ and increase ovarian cycle length.^80,85^ Additionally, Cd has also been found to affect follicle maturation, induce luteoolysis,^85,94^ and thicken endometrium.^23,85^ All these results suggest that exposure to Cd may impair women’s fertility. However, results obtained from epidemiological studies have been conflicting. Several cohort studies investigating associations between exposure to Cd and women’s fertility had conflicting results including no associations^95^ or even reduced fecundity.^96^ In contrast, Cd has also been found to disrupt reproductive hormone secretion.^78,97^ A study from Lee et al. discovered an inverse relationship between blood concentrations of Cd and Anti-Mullerian hormone (AMH) – a peptide hormone secreted from growing follicles and is commonly used as a biomarker of ovarian reserve, suggesting that exposure to Cd may increase women’s infertility risk by diminishing ovarian reserve.^98^ Collectively, as we study the role of nutrition status on the toxicokinetics of Cd, it is essential to integrate both experimental and epidemiological evidence and include all possible confounding factors to determine the effects of Cd on women’s reproductive health and fertility.

Experimental evidence reveals that Hg accumulates in the ovaries and impacts female reproduction^77,85,99^ by interfering with the secretion patterns of gonadotropins of LH and FSH, altering ovarian cyclicity, and inducing follicular cell apoptosis and follicle atresia.^85,100-102^ Although some other studies reported that Hg is associated with female infertility, the evidence to support this is limited and inconclusive.^77,85,103,104^ Thus, evidence is inadequate to draw meaningful conclusions about how Hg impacts female reproductive outcomes, underscoring the need for additional research.

### Heavy metal mixtures on women’s fertility in epidemiological and experimental studies

Previous studies have examined heavy metals and individual reproductive outcomes without examining the complexities of reproductive cycles and the interactions of these exposures. Both epidemiological and experimental literature is lacking for assessing the mixture of heavy metals on women’s reproductive outcomes. Previous studies assessing other health outcomes have used the simple concentration additive method or other statistical methods^105^ for combining metals.^39,40^ Here, we integrated multiple heavy metal concentrations by considering each individual metal’s toxicity related to the ER stress, a key mediator of the adverse outcome pathway in female reproduction.^49,106^ EPA’s framework for metal risk assessment outlines that some metals act additively while others are antagonistic or synergistic when they are present together.^107^ These interactions occur during absorption, excretion, or sequestration.^107^ However, the exact fate and joint effects of Pb, Cd, and Hg together in women has yet to be determined; additionally, metal mixtures in women can be dependent on other factors that are different across individuals, making it hard to quantify.

### The link between women’s infertility and long-term amenorrhea

The menstrual cycle, or periodic vaginal bleeding due to the shedding of uterine endometrium, is regulated by the cyclic changes of reproductive hormones, including both gonadotropins from the pituitary and sex hormones from the ovaries.^108^ Pathological amenorrhea that are not caused by pregnancy, lactation, or menopause occurs in 3 – 4% of women in the US.^109,110^ In our study, we found no association between women’s ever-infertility and their recent long-term amenorrhea, suggesting that women’s recent menstrual cycle status does not reflect their fertility history. It is also likely that amenorrhea is only one of many complex contributing factors towards women’s fertility success. For example, although up to 25% of infertile women have disturbed menstrual cycle such as amenorrhea,^1,111^ infertility can also be attributed to sperm defects from the male partner and other unexplained reasons.^4^ The underlying mechanism of women’s amenorrhea remains poorly understood and has been attributed to both genetic and environmental factors.^112,113^ In addition to causing infertility, amenorrhea can have additional health consequences. For example, continual anovulation for two to three years increases the risk of developing endometrial cancer,^114^ suggesting that long-term amenorrhea is a risk factor of other female reproductive disorders.

### Advantages and limitations

This study overcomes several limitations in previous papers using NHANES database to investigate associations between heavy metals and women’s infertility.^27,74^ First, both of these studies only included participants from two NHANES cycles (2013-2014 and 2015-2016), whereas we further added the cycle of 2017-2018. Second, the study from Lee et al. only included infertile women up to age 39 and compared them to pregnant women, which resulted in a smaller sample size of n=124.^27^ Here, in addition to women of 20-39 years, we also included women of 40-49 years, because these women may have experienced infertility before and are also within reproductive age; moreover, we defined the fertile women in two ways, including self-reported fertile women and pregnant women. Third, our study accounts for additional covariates related to reproduction that are essential for understanding infertility, such as hormonal contraception use, menstruation patterns, and possible help from a doctor for fertility issues; further, our study also examined the difference in infertility status among women who may have seen a doctor and gotten assistance to become pregnant vs. those who did not. Fourth, we chose to define fertile women in two sub-groups due to some limitations in the NHANES survey questions. The main-group or women who self-reported to be fertile or ever-infertile could include women who have never ‘tried’ to become pregnant. Therefore, it is possible to include several misclassifications. Thus, we additionally looked at a sub-group of current pregnant women. There were ten women who were pregnant but also answered “yes” to the question of “Have you ever attempted to become pregnant over a period of at least a year without becoming pregnant?” We chose to include these ten women in the ever-infertile group because they reported having had issues with their fertility in the past. Indeed, five of those ten women responded that they had previously seen a doctor because they were unable to become pregnant, indicating that these women likely have received ART such as IVF to become pregnant. Lastly, this study takes a unique approach to assessing the reproductive toxicity of the mixture of heavy metals using weighted TEQ values.

Our study has several limitations due to NAHNES study design, the complexities of assessing female reproduction and reproductive toxicities of heavy metals. First, there are limitations due to NHANES study design. NHANES is a cross-sectional study, therefore casual and temporal relationships cannot be confirmed. Cross-sectional data are also prone to survival bias. Although women aged 15-19 are still considered of reproductive age, we did not include them because the study design did not collect exposure or outcome information in this age group. Additionally, NHANES did not collect information on some reproductive diseases that also play a role in infertility, such as endometriosis and PCOS. For example, about 30 to 50% of women with endometriosis are infertile and PCOS is a leading cause of infertility.^115,116^ Moreover, the NHANES questionnaire only collected historical use of birth control and female hormones. Second, the reproductive health outcomes are measured using a self-reported questionnaire. Although self-reported information is useful, various definitions may affect the prevalence of a measured outcome. With the information collected, we did our best to define the outcomes (ever-infertile, fertile, pregnant, long-term amenorrhea, menstruating). However, there are limitations for the definitions we used. For example, amenorrhea is defined as the absence of menstruation for at least a 90-day period.^117^ However, NHANES only collects information on absence of menstruation for the past 12 months.^57^ Although menopause can also be defined by one year of no menses, we chose to name our variable long-term amenorrhea because these women self-reported not having menopause. Thus, women categorized with long-term amenorrhea may have suspected early menopause or POF. Third, male factors account for approximately 40-50% of all cases of infertility.^118^ The NHANES questionnaire only addressed females. Therefore, male infertility factors were not considered. However, the relationship between male reproduction and heavy metals has been well studied, while female reproductive function is lacking.

Understanding what blood metal concentrations represent is also worth discussing. A single measurement of blood metal concentration may not reflect long-term exposure though some studies suggested that under steady state conditions a single measurement of blood metal level seems to be acceptable as it can reflect body metal burden of long-term exposure.^119^ Our study assumes women’s blood metal concentrations during the time of the examination were the same as when they experienced infertility or long-term amenorrhea. However, by study design, there is no way of knowing temporality. Although the biological half-lives for heavy metals in the human body are long, the half-lives in blood specifically can be shorter and vary (Hg = 50 days,^120^ Cd = 3-4 months for the fast component and 7-16 years for the slow component,^121-123^ Pb = 1-2 months^124^). Blood metal concentrations are used to represent both recent and chronic exposures.^125,126^ However, it is important to note that the acute exposures can modify blood metal concentrations. For example, eating fish right before the examination could markedly elevate blood Hg concentrations, however, someone who has been chronically exposed to Hg, maintains high concentrations in their blood even after exposure has ended.^127,128^ This same concept could be applied to Cd blood concentrations with a participant who smoked before the examination. Our study assumes that the individual’s behavior prior to the examination is consistent to their daily behaviors. Additionally, those with chronic past exposure are often underestimated when assessing blood levels because metals like Pb can be stored in the bone. Therefore, individuals can have a high body burden of Pb but still appear to have normal Pb concentrations in the blood.^129^ Lastly, we used the ER stress, an important contributing factors of female reproductive dysfunctions, to calculate the mixture score of heavy metals because the Tox21 program has the screening results of all three metals available. However, it is possible that heavy metals may compromise female reproduction through other mechanisms such as DNA damage, oxidative stress, and epigenetic modification. Thus, an optimized calculation method of heavy meatal mixtures is highly desired.

## Conclusion

In summary, the results of our studies using NHANES 2013-2018 reveal that there are significant percentages of women having blood heavy metal levels exceeding typical or normal levels. Moreover, the blood concentrations of single Pb and heavy metal mixtures are associated with an increase of women’s historical infertility. This study highlights the threat of heavy metal exposure on women’s reproductive health and fertility as well as an urgent unmet need to prevent and reduce heavy metal exposure.

## Data Availability

All data were obtained from NHANES, a nationally representative cross-sectional survey of the non-institutionalized U.S. population. NHANES is conducted by the US Centers for Disease Control and Prevention (CDC) and uses a complex multistage, probability sampling design.

## Authors’ roles

M. McClam, J. Liu, and S. Xiao conceived of the project, contributed to the experimental design, data collection and analysis, data interpretation, and manuscript writing. Y. Fan and T. Zhan contributed to the data collection and analysis. Q. Zhang, DE. Porter, and GI. Scott contributed to the data interpretation and manuscript writing.

## Funding and acknowledgments

This work was supported by the National Institutes of Health (NIH) K01ES030014, P30ES005022 and UH3ES029073 to S. Xiao, R01ES032144 to S. Xiao and Q. Zhang, P01ES028942 to GI. Scott, D. Porter, and S. Xiao, and start-up fund from the Environmental and Occupational Health Sciences Institute (EOHSI) at Rutgers University to S. Xiao.

